# Automated Evaluation of Large Language Model Response Concordance with Human Specialist Responses on Physician-to-Physician eConsult Cases

**DOI:** 10.1101/2025.08.14.25332839

**Authors:** David JH Wu, Fateme Nateghi Haredasht, David Wu, Vishnu Ravi, Liam G. McCoy, Yingjie Weng, Kanav Chopra, Selin S. Everett, George Nageeb, Wenyuan Chen, Stephen P. Ma, Saloni Kumar Maharaj, Jessica Tran, Leah Rosengaus, Lena Giang, Olivia Jee, Ethan Goh, Jonathan H Chen

## Abstract

Specialist consults in primary care and inpatient settings typically address complex clinical questions beyond standard guidelines. eConsults have been developed as a way for specialist physicians to review cases asynchronously and provide clinical answers without a formal patient encounter. Meanwhile, large language models (LLMs) have approached human-level performance on structured clinical tasks, but their real-world effectiveness requires evaluation, which is bottlenecked by time-intensive manual physician review. To address this, we evaluate two automated methods: LLM-as-judge and a decompose-then-verify framework that breaks down AI answers into verifiable claims against human eConsult responses. Using 40 real-world physician-to-physician eConsults, we compared AI-generated responses to human answers using both physician raters and automated tools. LLM-as-judge outperformed decompose-then-verify, achieving human-level concordance assessment with F1-score of 0.89 (95% CI: 0.750, 0.960) and Cohen’s kappa of 0.75 (95% CI 0.47,0.90) —comparable to physician inter-rater agreement κ = 0.69-0.90 (95% CI 0.43-1.0).

## 1. Introduction

Electronic consultations (eConsults) expand specialist access by enabling generalists to asynchronously obtain expert advice without an in-person referral. Real-world studies have demonstrated faster care and clinical quality comparable to face-to-face consults^1,2^. Despite growing adoption, eConsult programs often face workflow bottlenecks for both referring physicians and specialists, as formulating high-quality consults and responses requires retrieving dispersed clinical data, synthesizing relevant context, and drafting clear, actionable recommendations. These challenges have prompted growing interest in AI systems that can streamline and augment key steps in the consultation workflow. Early pilot use cases of large language models (LLMs) reduce inbox burden and documentation time^3–5^ and now match or surpass clinicians on standardized medical vignettes^4,6–12^ and, when paired with physicians, can boost diagnostic accuracy^13–15^.

Stanford Health Care operates a mature multispecialty eConsult program that generates a retrospective corpus of real physician-to-specialist consults. This dataset includes question-and-answer pairs with accompanying clinical context—offering a valuable resource for evaluating AI systems in realistic workflows. As part of ongoing efforts to augment end-to-end eConsult workflows, we have been developing SAGE (Specialist AI Guiding Experts), a system combining vector embeddings, predictive models, and generative AI to route clinical queries to the correct specialty, retrieve relevant chart information, suggest evidence-backed recommendations, and draft specialist-level replies to physician queries.

Recent LLM evaluation studies rely on polished vignettes or synthetic benchmarks that sanitize the inherent ambiguity and incompleteness of real-world cases^16,17^. Real-world perturbations, such as changes in geography, patient demographics, idiosyncratic phrasing, or missing context, can swing diagnostic accuracy by 20 percentage points^18^, suggesting current benchmarks overestimate clinical readiness. Moreover, human evaluation of LLM outputs is labor-intensive, costly, and difficult to scale. Researchers are responding by developing more comprehensive benchmarks to evaluate their performance^19–23^.

In this study, we compare two scalable, LLM-assisted evaluation methods against human specialist ratings on a diverse set of real eConsults: (1) an LLM-as-judge approach directly scores concordance between SAGE drafts and human responses; (2) a decompose-then-verify framework, inspired by and adapted from current existing methods MedScore, VeriFact, which breaks AI outputs into atomic claims for fact-checking^24,25^. We benchmark both approaches against blinded ratings from board-certified specialists to determine whether LLM-assisted grading can reliably and efficiently approximate human judgment when comparing AI versus expert concordance. By assessing robustness across multiple specialties and noisy, unstructured cases, we ask whether these methods provide reliable, low-cost grading that can keep pace with rapid model iterations, ultimately enabling safer, large-scale deployment of LLM-augmented eConsult workflows.

## 2. Methods

### 2.1 Data Collection and Study Population

A random sampling of 50 physician-to-physician eConsult pairs performed from October 2022 to March 2024 was collected from Stanford Health Care’s clinical data. The eConsult program was initiated as a way for general physicians to consult advanced specialties such as dermatology and hematology virtually via the EHR. Relevant consult notes were extracted from the EHR using structured metadata (e.g., specialty, consult type, and date ranges) and keyword-matched section headers (e.g., “eConsult Question” and “eConsult Response”). All data were de-identified using the Safe Harbor method according to National Institute of Standards and Technology (NIST) guidelines, and the clinical text was further anonymized using the TiDE algorithm to ensure compliance with privacy regulations. The answering physicians in eConsult provided clinical recommendations based solely on chart review and did not see the patient or perform a physical examination. Based on physician discretion, some patients would be deemed medically appropriate for referral to their respective specialty clinic. The eConsult program did not permit back-and-forth conversation between physicians. If the consulting physician’s question could not be answered satisfactorily, the patient was referred to the specialty clinic for further evaluation, or the consult was declined due to reasons such as the wrong scope of specialty or lack of clarifying details. The eConsult consult-and-answer pairs were then de-identified again using a BERT-based method and manually reviewed by DW, DJW to confirm removal of any identifying information. Duplicate, incomplete, or erroneous consults were excluded from the study. This study was conducted with IRB approval (IRB-47618) to ensure compliance with ethical research standards.

### 2.2 AI eConsult Response Generation

AI responses were generated using a secure, HIPAA-compliant large language model instance (“SecureGPT”) approved for PHI analysis at Stanford University, utilizing OpenAI GPT-4.1 as the underlying model. A standardized prompt was developed to structure AI responses into Assessment, Recommendations and Rationale, Contingency Plan, and Citations sections. To provide appropriate clinical context, the three most recent clinical Progress Note with length >3,500 characters (we found in initial pilot work that those less than this cut-off were more likely to be clerical notes without meaningful clinical significance), prioritizing the consulted specialty in question, were also provided in the API query, as well as the 20 most recent lab values.

**Table 1.**
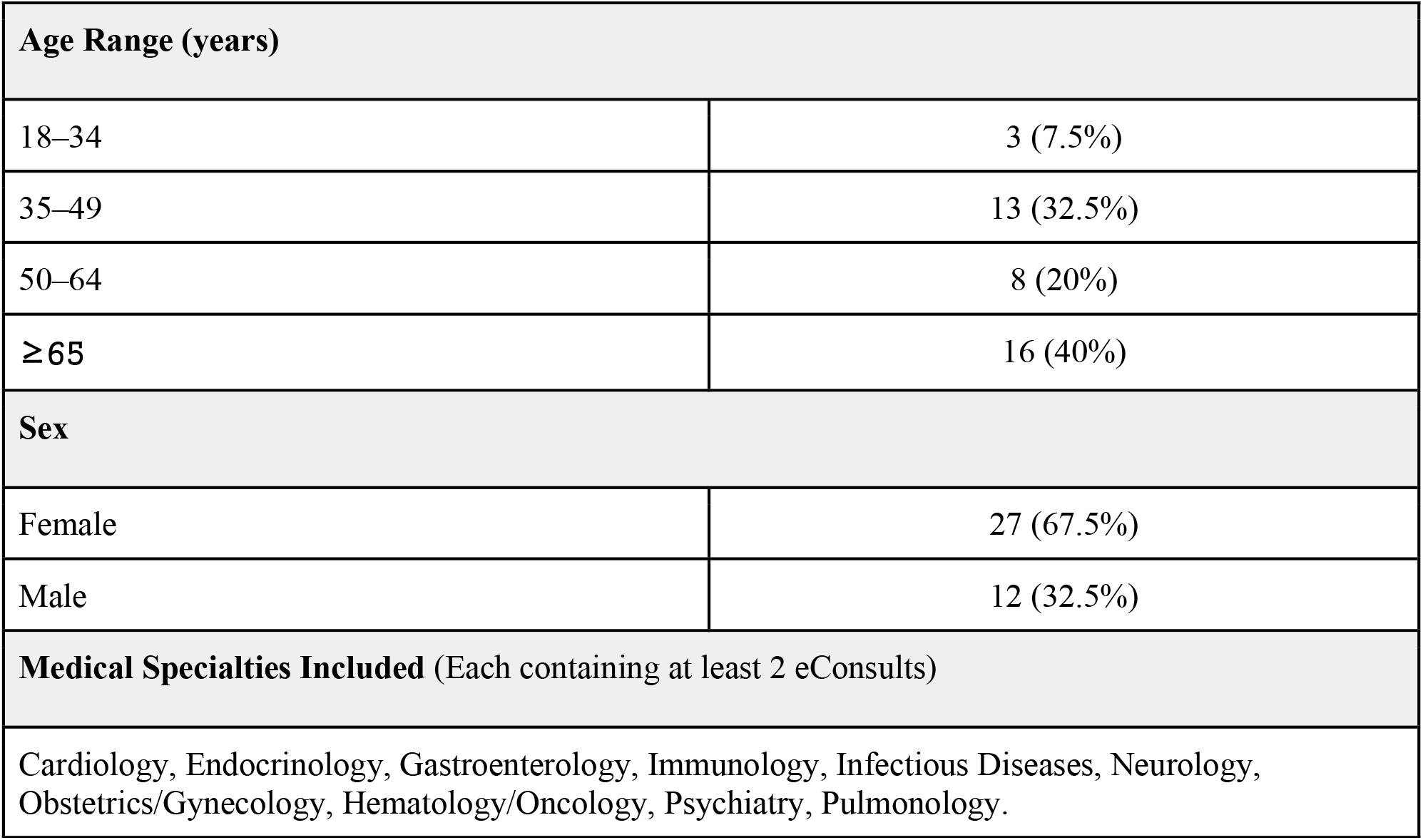
Patient eConsult demographics by consult type, age, and sex.

### 2.3 LLM-assisted Concordance Evaluation

To assess the concordance between AI-generated and human-generated eConsult responses, we developed two LLM-assisted evaluation workflows, with the original specialist consult answer as a “silver-label answer”. For our two LLM-assisted concordance evaluation workflows, one was based on using *LLM-as-Judge (LaJ)* and the other using a *Decompose-then-Verify* (*DtV)* approach.

In our *LaJ* approach, we provided a concordant and discordant example case not included in the data set to provide as a reference in the prompt. API calls were made using a secure, HIPAA-compliant, and PHI-safe institution-specific endpoint.

In our *DtV* approach, we implemented a two-stage pipeline adapted from and inspired by the MedScore and VeriFact frameworks^22,23^. First, AI responses were decomposed into atomic, independently verifiable medical claims using a specialized LLM query. Second, each atomic claim was verified against the original human specialist response using a separate verification query. Claims with ≥80% support percentage were classified as concordant. The pipeline was implemented in Python. Three of the authors (DJW, DW, LM) not involved in the final physician review reviewed 4 examples of cases decomposed into atomic claims and their corresponding LLM-assisted verification. Consensus determination was that overall performance was satisfactory, and a sampling of these cases was included as examples in both the decomposition and the verification prompt.

### 2.4 Human Physician Evaluation

Forty eConsult question-and-answer pairs were provided to three blinded attending internal medicine attending physicians (VS, SM, JT) for concordance evaluation. Physicians rated concordance between AI-generated and human specialist responses on a binary scale (1 = Concordant, 0 = Discordant). For concordant cases, raters indicated their preference between the AI and human responses. For equipoise, only cases in which AI and human were rated as concordant were used in Preference rating. Prior to rating, the physician reviewers participated in a calibration session to establish consensus on concordance definitions and rating criteria.

### 2.5 Statistical Analysis

Aggregate physician concordance ratings were determined by majority vote (best-of-three). Inter-rater agreement among the three physician evaluators was calculated using Fleiss’ kappa for multi-rater categorical data. Agreement between physician ratings and LLM-assisted concordance evaluations was assessed using Cohen’s kappa for pairwise comparisons. Kappa values were interpreted as: <0.20 (poor), 0.21-0.40 (fair), 0.41-0.60 (moderate), 0.61-0.80 (substantial), and 0.81-1.00 (almost perfect) agreement.

Performance metrics for both LLM evaluation approaches were calculated, including sensitivity, specificity, precision, recall, and F1-score (calculated as harmonic mean of precision and recall) using physician majority vote as the reference standard. Ninety-five percent confidence intervals for all kappa and F1 coefficients were calculated using bias-corrected and accelerated (BCa) bootstrap resampling. The BCa method accounts for both bias and skewness in the bootstrap distribution, providing more accurate coverage than standard percentile methods. All statistical analyses were performed using Python with scikit-learn and statsmodels libraries.

## 3. Results

### 3.1 LLM-Assisted Concordance Evaluation Performance

Forty eConsult cases were included in the final analysis after excluding duplicate, incomplete, or erroneous eConsults, with an equal distribution of 20 concordant and 20 discordant cases based on physician majority vote. Both LLM evaluation approaches demonstrated substantial performance in concordance assessment, with LaJ consistently outperforming the DtV approach (Figure 2). The DtV approach achieved moderate performance (F1-score: 0.73 [95% CI: 0.55, 0.86], κ = 0.40, [95% CI: 0.12, 0.65]), while LaJ methods showed substantial to near-perfect agreement. Among LaJ implementations, DeepSeek R1 achieved the highest performance (F1-score: 0.89 [95% CI: 0.750, 0.960], κ = 0.75, [95% CI: 0.47, 0.90]), followed by Gemini 2.5 Pro (F1-score: 0.86 [95% CI: 0.44, 0.89], κ = 0.70, [95% CI: 0.44, 0.89]). All LaJ approaches demonstrated superior concordance evaluation compared to the claim-based DtV method.

**Figure 1.**
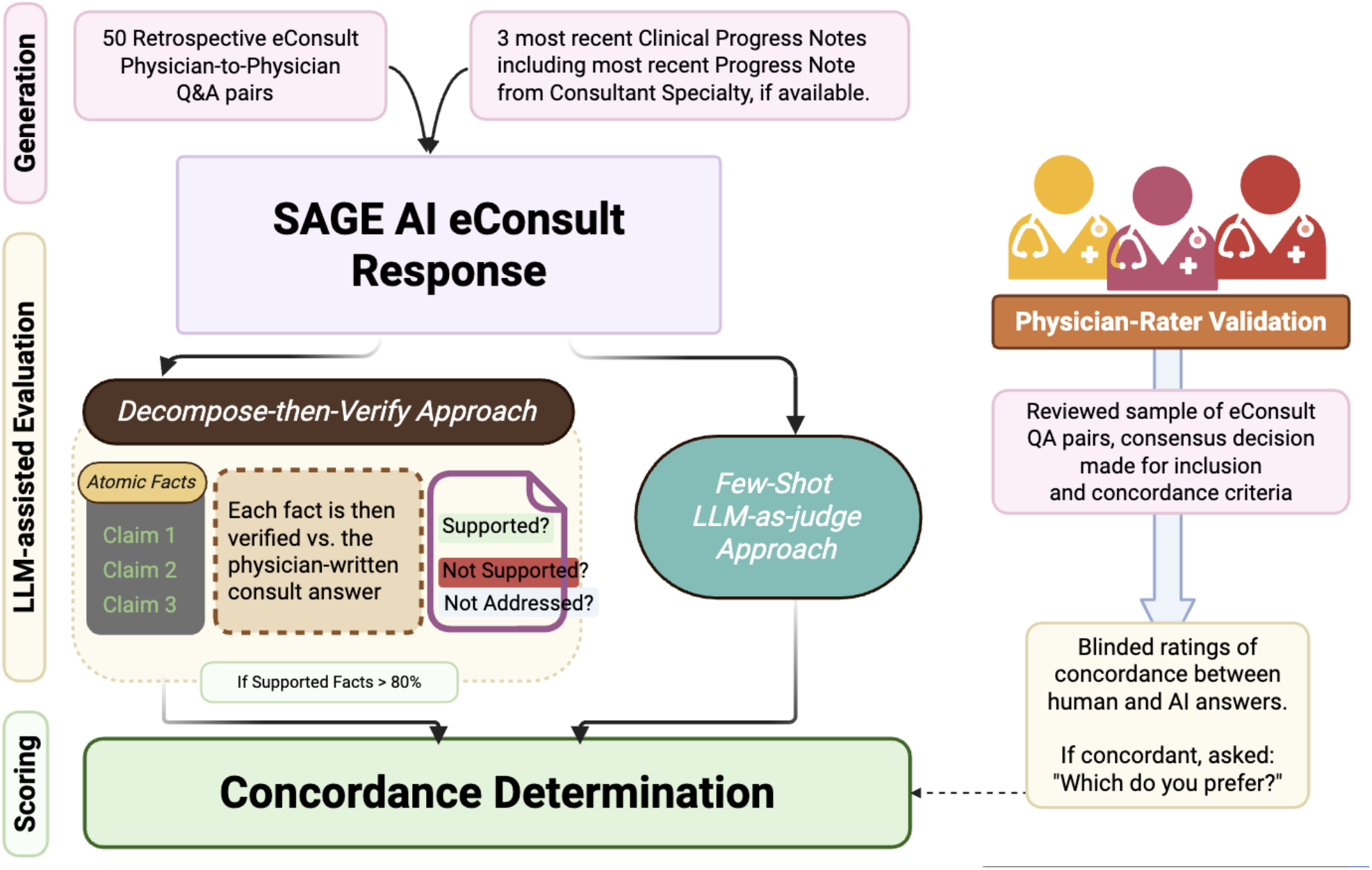
Schematic of study design. 50 real-world retrospective eConsult cases were collected. We provided the original consult question as well as clinical context consisting of the three most recent clinical progress notes and 20 most recent lab values, generating an AI e-Consult response. We then manually assessed concordance of the AI response to human response and also performed LLM-assisted automated concordance evaluations.

**Figure 2.**
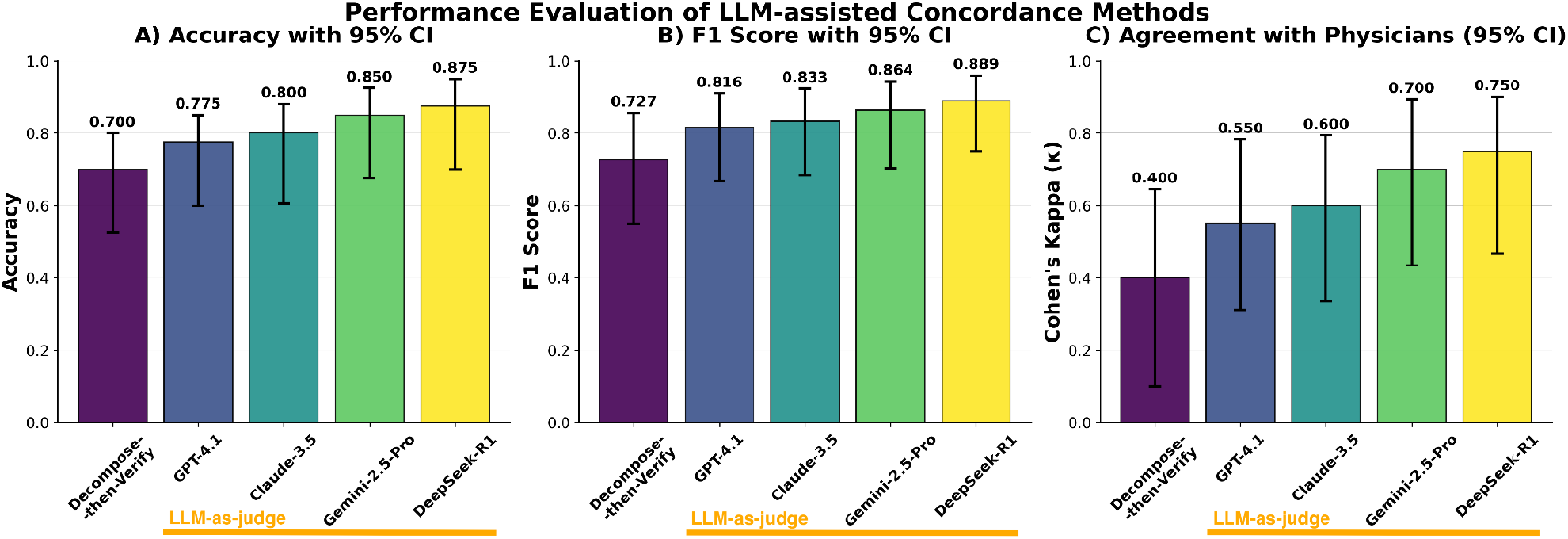
Three-panel comparison showing (A) Accuracy, (B) F1-score, and (C) Cohen’s kappa agreement with physician majority vote for five different approaches. LLM-as-Judge methods (GPT-4.1, Gemini 2.5 Pro, DeepSeek R1, Claude 3.5) consistently outperformed the Decompose-then-Verify approach across all metrics. DeepSeek R1 achieved the highest performance with near-human-level agreement (κ = 0.75, 95% CI 0.47-0.90)

The DtV pipeline decomposed AI responses into an average of 24.9 atomic claims per case (range: 17-34 claims). Of these claims, 41.0% were classified as “Supported,” 10.1% as “Not Supported,” and 48.9% as “Not Addressed” by the reference specialist consult answer, suggesting a significant amount of extra information added in the AI consult answers. ROC analysis of the support percentage threshold revealed an AUC of 0.745.

### 3.2 Physician and LLM Inter-Rater Concordance Agreement

Inter-rater agreement among the three physician evaluators was moderate (Fleiss’ κ = 0.456, n=32). Perfect agreement across all three raters occurred in 61.3% of cases, indicating variability in concordance assessment even among experienced clinicians. Individual physician agreement rates with the aggregate majority vote were substantial: Physician 1 (κ = 0.90, 95% CI 0.70-1.0), Physician 2 (κ = 0.69, 95% CI 0.43-0.88), and Physician 3 (κ = 0.69, 95% CI 0.42-0.89), with Physician 1 demonstrating the highest consistency with the consensus rating. Pairwise agreement between individual physicians showed moderate agreement: Physician 1 vs Physician 2 (κ = 0.57), Physician 1 vs Physician 3 (κ = 0.59), and Physician 2 vs Physician 3 (κ = 0.34). When comparing LLM-assisted methods to individual physician raters, several patterns emerged (Figure 3). DeepSeek R1 achieved substantial agreement with all three physicians (κ = 0.65, 0.61, 0.55), while Gemini 2.5 Pro demonstrated strong agreement with Physicians 1 and 2 (κ = 0.70, 0.61) but more moderate agreement with Physician 3 (κ = 0.44). The Decompose-then-Verify approach showed consistently moderate agreement across all physician raters (κ = 0.40, 0.30, 0.42). Notably, the best-performing LLM methods achieved agreement levels with individual physicians that were comparable to inter-physician agreement

**Figure 3.**
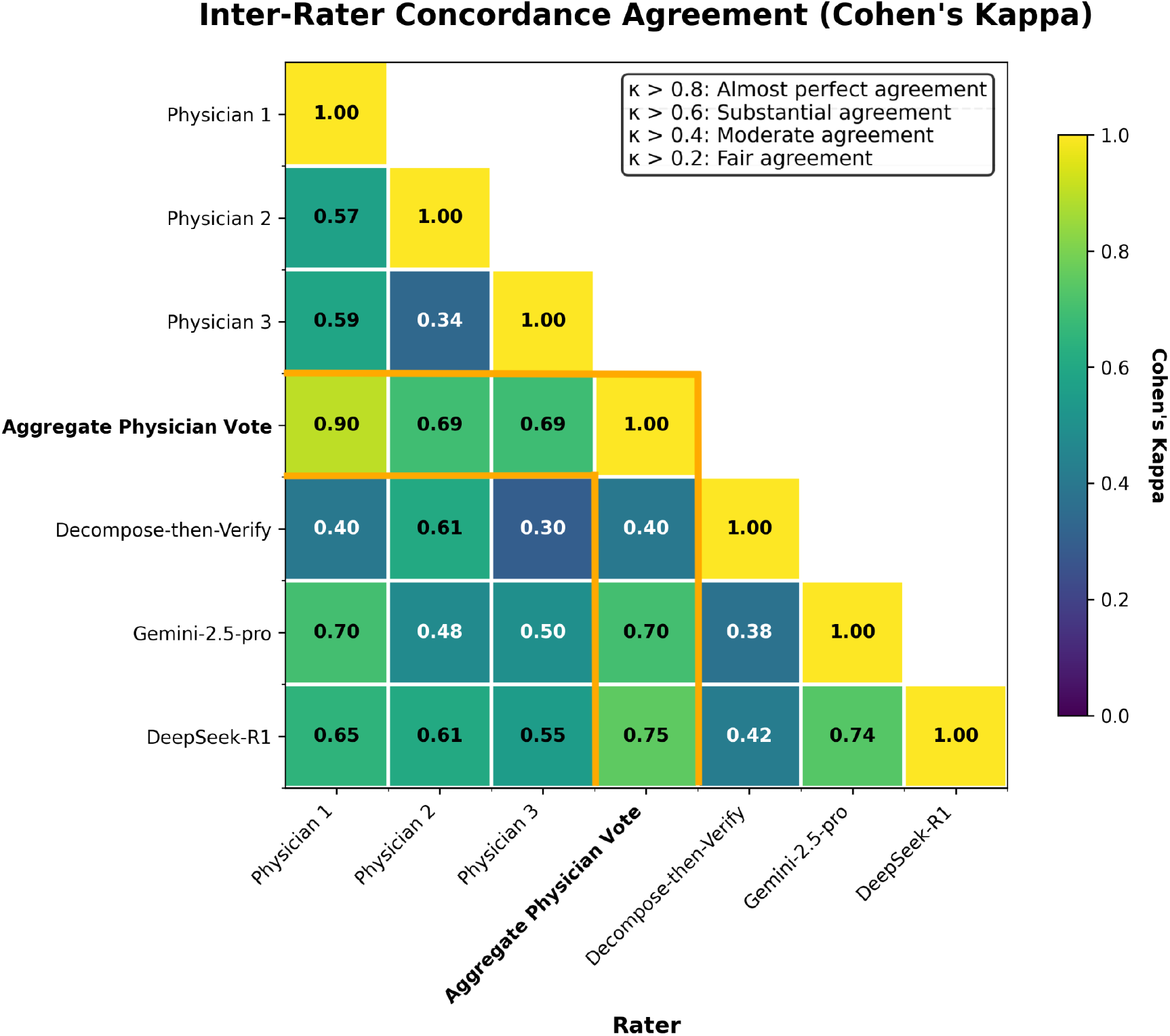
Inter-rater Agreement Heatmap for Concordance Evaluation. Cohen’s kappa coefficients between all rater pairs, displayed as a lower-triangular heatmap. Values range from 0 (no agreement) to 1 (perfect agreement), with darker colors indicating higher agreement. The aggregate physician vote represents majority consensus among the three physician raters. LLM-assisted methods (DeepSeek R1, Gemini 2.5 Pro) achieved substantial agreement with physician raters, comparable to inter-physician agreement levels.

### 3.3 Physician Preference Analysis

Among the 20 cases rated as concordant by majority vote, physician preferences between AI and human responses showed substantial heterogeneity (Figure 4). Individual preference patterns varied dramatically: Physician 1 preferred AI responses in 33.3% of concordant cases, Physician 3 in 12.5%, and Physician 2 in 81.8% of cases. This divergence in preferences highlights the subjective nature of response quality assessment, even when both AI and human responses are deemed clinically appropriate. Inter-rater agreement on preferences was poor (Fleiss’ κ = −0.12, n=11), with pairwise agreements ranging from κ = −0.23 (Physician 1 vs Physician 3) to κ = 0.19 (Physician 1 vs Physician 2). The negative kappa values indicate systematic disagreement beyond what would be expected by chance, suggesting that physicians may have intrinsic differences in preference and criteria for those preferences, even when responses are clinically concordant.

**Figure 4.**
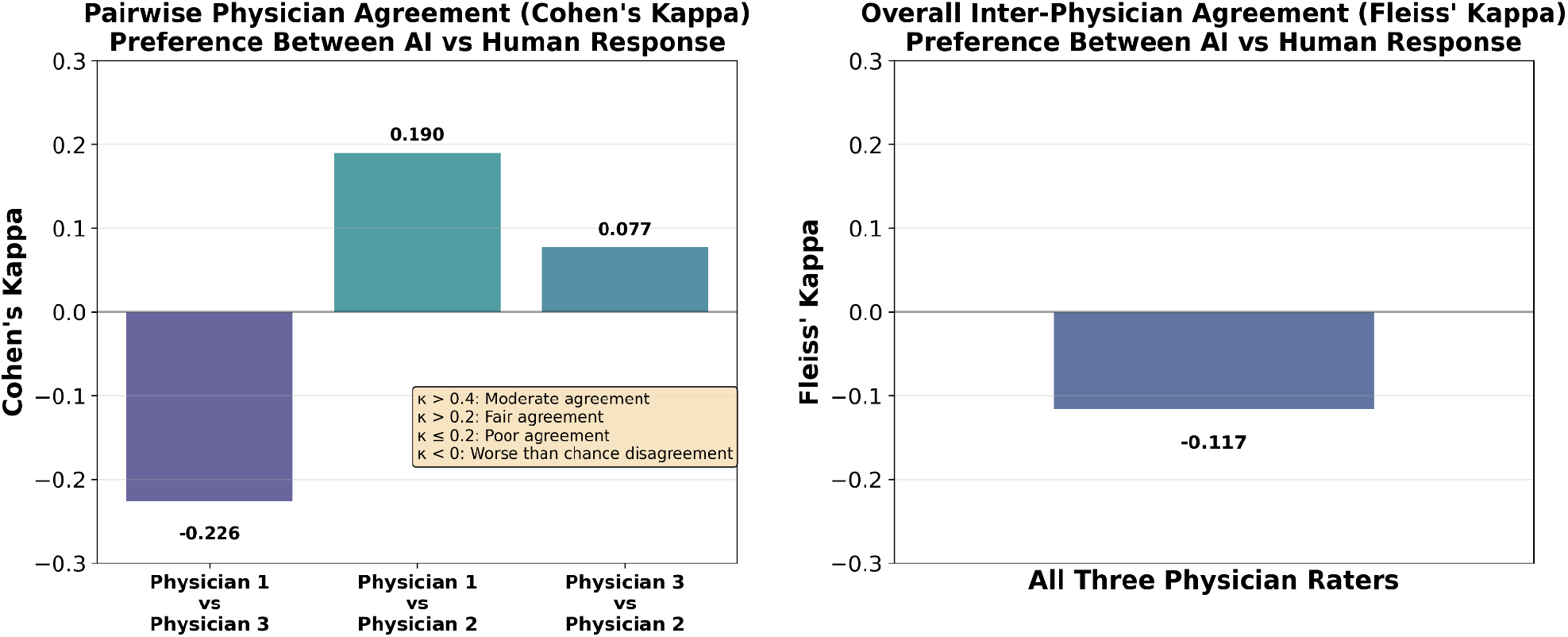
(A) Pairwise Cohen’s kappa coefficients between physicians for preference assessment (AI vs human responses), showing poor agreement with negative values indicating systematic disagreement. (B) Overall, Fleiss’ kappa (κ = −0.12) demonstrates poor multi-rater agreement on preferences. Analysis limited to the 20 cases where all three physicians agreed responses were clinically concordant, highlighting that clinical acceptability does not predict preference uniformity

**Figure 5.**
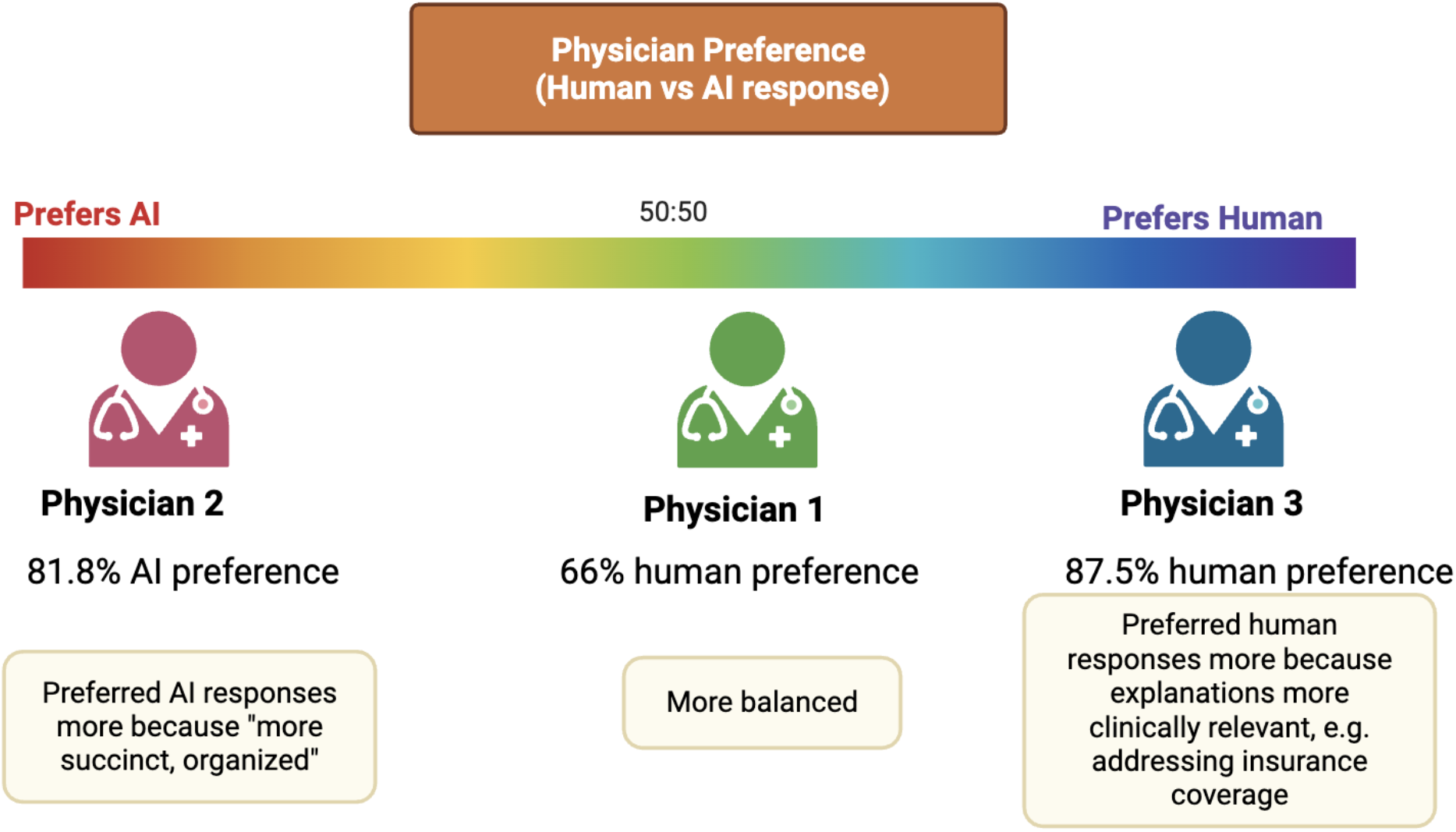
Physician Preference Distribution of Clinically Concordant Human vs AI responses. *Significant differences in patterns of preference were observed between our physician raters*.

## 4. Discussion

This study demonstrates that LLM-assisted evaluation of concordance between AI-generated and human specialist eConsult responses achieves substantial performance comparable to inter-physician agreement. The LLM-as-Judge approach consistently outperformed the Decompose-then-Verify method, with the best-performing model (DeepSeek R1) achieving an F1-score of 0.89 (95% CI: 0.75, 0.96) and Cohen’s κ of 0.75 (95% CI: 0.47, 0.90)—approaching the range of inter-physician agreement (κ = 0.69-0.90, 95% CI: 0.43, 1.0) observed in clinical settings. This represents the first validation of automated concordance evaluation in real-world physician-to-physician eConsultations, addressing a critical bottleneck in scalable LLM evaluation for clinical applications.

Our findings address a key challenge in clinical AI evaluation: the labor-intensive nature of expert review that limits scalable assessment of LLM performance in real-world settings. Traditional evaluations rely heavily on standardized clinical vignettes, which may overestimate performance in practice due to their structured nature and absence of real-world ambiguity. By demonstrating that LLM-assisted evaluation can reliably assess concordance in unstructured eConsult cases, this work provides a pathway for large-scale retrospective benchmarking of clinical LLMs prior to deployment that can be tailored to each institution.

Interestingly, we noted substantial heterogeneity in physician preferences (Fleiss’ κ = −0.12) even among concordant cases, highlighting an important distinction between clinical acceptability and physician preference: while responses may be equally acceptable from a clinical standpoint, individual physicians maintain distinct preferences regarding communication style, comprehensiveness, and presentation format. One physician rater noted that the human responses took into account factors such as insurance or even manually reviewed EKG findings themselves to provide their own insight, which is beyond the capabilities of the current AI eConsult system. The substantial preference heterogeneity suggests that AI integration in clinical practice may benefit from personalization approaches and further iterative improvement that accommodate individual physician preferences while maintaining clinical appropriateness.

The superior performance of LaJ over DtV approaches likely reflects the complexity of medical reasoning that is not entirely captured by simple decomposition and verification of atomic claims. Clinical decision-making often involves nuanced integration of multiple factors, contextual considerations, and implicit medical knowledge that may be lost when responses are parsed into discrete verifiable statements. The DtV approach’s moderate performance suggests utility for systematic claim verification but indicates limitations for holistic clinical assessment. Interestingly, 48.9% of all claims generated were classified as ‘Not Addressed’ by the reference material, suggesting that AI responses often included extra details that the human physician did not. In discussion with the physician raters, it was usually only one or two key “atomic claims” in each case, such as the decision to start a new drug or not, that pushed them to decide whether a case was concordant or discordant, whereas in the DtV approach each claim is weighted equally relative to the other claims.

Several limitations warrant consideration. The sample of 40 cases evaluated is not intended to generalize across specialties and clinical scenarios, but was sufficiently powered to confirm the potential for this methodology to systematically evaluate a volume of cases that would be impractical for manual review. Human specialist responses are used here as a comparison standard for comparison, but this should NOT be used to assume that actual care delivered represents optimal care decisions. Existing evidence and standards of care establish a high correlation between actual and preferred clinical decision-making, while also acknowledging legitimate and acceptable variations in clinical approaches. Our evaluation focused on concordance rather than clinical outcomes, leaving questions about whether concordant responses lead to better patient care. Concordance with human experts is thus not the only critical measure of medical advice, but an important one, especially in complex clinical scenarios where “correct” answers are often poorly defined with limited definitive evidence. It is indeed often the case where limited evidence or guidelines are available that physicians reach for specialist consultation.

During the process of analysis, we also found in discussion with our physician reviewers that concordance is difficult to rate in a binary fashion, there is often a spectrum of concordance, with some cases being definitely concordant while others fall in a gray zone; sometimes answers are concordant in their recommended intervention but differ in their contingency plans. This complexity in concordance rating likely explains the moderate inter-rater agreement among physicians, highlighting the nuances of medical reasoning and decision making that complicates concrete ground truth for clinical evaluations. We originally anticipated that the systematic decomposition and then claim-by-claim verification of the DtV method would better capture these nuances but were ultimately surprised by the superior and human-level performance of the LaJ approach, hinting at the rapid improvement of these models’ innate clinical reasoning abilities on a clinical concordance determination task that is difficult even for licensed physicians.

## Conclusion

This study demonstrates that large language models can be evaluated on real-world physician-to-physician eConsults using automated concordance methods, with LLM-as-Judge (LaJ) approaches achieving performance comparable to human physician raters. Compared to decomposition-based verification methods, LaJ more reliably captured overall clinical concordance between AI- and human-generated responses.

## Data Availability

All code produced in the present study are available upon reasonable request to the authors

## Bibliography

1. Palen TE, Price D, Shetterly S, Wallace KB. Comparing virtual consults to traditional consults using an electronic health record: an observational case–control study. BMC Medical Informatics and Decision Making. 2012;12(1):65. doi:10.1186/1472-6947-12-65

2. Peeters KMM, Reichel LAM, Muris DMJ, Cals JWL. Family Physician–to–Hospital Specialist Electronic Consultation and Access to Hospital Care: A Systematic Review. JAMA Network Open. 2024;7(1):e2351623. doi:10.1001/jamanetworkopen.2023.51623

3. Garcia P, Ma SP, Shah S, et al. Artificial Intelligence-Generated Draft Replies to Patient Inbox Messages. JAMA Netw Open. 2024;7(3):e243201. doi:10.1001/jamanetworkopen.2024.3201

4. Hirosawa T, Kawamura R, Harada Y, et al. ChatGPT-Generated Differential Diagnosis Lists for Complex Case-Derived Clinical Vignettes: Diagnostic Accuracy Evaluation. JMIR Med Inform. 2023;11:e48808. doi:10.2196/48808

5. Wu DJ, Bibault JE. Pilot applications of GPT-4 in radiation oncology: Summarizing patient symptom intake and targeted chatbot applications. Radiotherapy and Oncology. 2024;190. doi:10.1016/j.radonc.2023.109978

6. Rutledge GW. Diagnostic accuracy of GPT-4 on common clinical scenarios and challenging cases. Learn Health Syst. 2024;8(3):e10438. doi:10.1002/lrh2.10438

7. Rao A, Pang M, Kim J, et al. Assessing the Utility of ChatGPT Throughout the Entire Clinical Workflow: Development and Usability Study. Journal of Medical Internet Research. 2023;25(1):e48659. doi:10.2196/48659

8. Yavuz YE, Kahraman F. Evaluation of the prediagnosis and management of ChatGPT-4.0 in clinical cases in cardiology. Future Cardiol. 20(4):197–207. doi:10.1080/14796678.2024.2348898

9. Greif C, Mpunga N, Koopman IV, Pye A, Hivnor CM, Owen JL. Evaluating the effectiveness of ChatGPT4 in the diagnosis and workup of dermatologic conditions. Dermatol Online J. 2024;30(4). doi:10.5070/D330464104

10. Mikhail D, Milad D, Antaki F, et al. Multimodal Performance of GPT-4 in Complex Ophthalmology Cases. J Pers Med. 2025;15(4):160. doi:10.3390/jpm15040160

11. Galetta K, Meltzer E. Does GPT-4 have neurophobia? Localization and diagnostic accuracy of an artificial intelligence-powered chatbot in clinical vignettes. Journal of the Neurological Sciences. 2023;453:120804. doi:10.1016/j.jns.2023.120804

12. Brodeur PG, Buckley TA, Kanjee Z, et al. Superhuman performance of a large language model on the reasoning tasks of a physician. Published online June 2, 2025. doi:10.48550/arXiv.2412.10849

13. Goh E, Gallo RJ, Strong E, et al. GPT-4 assistance for improvement of physician performance on patient care tasks: a randomized controlled trial. Nat Med. 2025;31(4):1233–1238. doi:10.1038/s41591-024-03456-y

14. Goh E, Gallo R, Hom J, et al. Large Language Model Influence on Diagnostic Reasoning: A Randomized Clinical Trial. JAMA Network Open. 2024;7(10):e2440969. doi:10.1001/jamanetworkopen.2024.40969

15. Jabbour S, Fouhey D, Shepard S, et al. Measuring the Impact of AI in the Diagnosis of Hospitalized Patients: A Randomized Clinical Vignette Survey Study. JAMA. 2023;330(23):2275–2284. doi:10.1001/jama.2023.22295

16. Bedi S, Liu Y, Orr-Ewing L, et al. Testing and Evaluation of Health Care Applications of Large Language Models: A Systematic Review. JAMA. 2025;333(4):319–328. doi:10.1001/jama.2024.21700

17. Evaluation and mitigation of the limitations of large language models in clinical decision-making | Nature Medicine. Accessed July 31, 2025. https://www.nature.com/articles/s41591-024-03097-1

18. Asiedu M, Tomasev N, Ghate C, et al. Contextual Evaluation of Large Language Models for Classifying Tropical and Infectious Diseases. Published online January 15, 2025. doi:10.48550/arXiv.2409.09201

19. Bedi S, Cui H, Fuentes M, et al. MedHELM: Holistic Evaluation of Large Language Models for Medical Tasks. Published online June 2, 2025. doi:10.48550/arXiv.2505.23802

20. Ma MD, Ye C, Yan Y, et al. CliBench: A Multifaceted and Multigranular Evaluation of Large Language Models for Clinical Decision Making. Published online October 11, 2024. doi:10.48550/arXiv.2406.09923

21. Wu C, Qiu P, Liu J, et al. Towards evaluating and building versatile large language models for medicine. npj Digit Med. 2025;8(1):1–13. doi:10.1038/s41746-024-01390-4

22. Arora RK, Wei J, Hicks RS, et al. HealthBench: Evaluating Large Language Models Towards Improved Human Health.

23. McCoy LG, Swamy R, Sagar N, et al. Do Language Models Think Like Doctors? Published online February 12, 2025:2025.02.11.25321822. doi:10.1101/2025.02.11.25321822

24. Chung P, Swaminathan A, Goodell AJ, et al. VeriFact: Verifying Facts in LLM-Generated Clinical Text with Electronic Health Records. Published online January 28, 2025. doi:10.48550/arXiv.2501.16672

25. Huang H, DeLucia A, Tiyyala VM, Dredze M. MedScore: Factuality Evaluation of Free-Form Medical Answers. Published online May 24, 2025. doi:10.48550/arXiv.2505.18452

